# Functional annotation of the human PTSD methylome identifies tissue-specific epigenetic variation across subcortical brain regions

**DOI:** 10.1101/2023.04.18.23288704

**Authors:** Hongyu Li, Jiawei Wang, Dianne A. Cruz, Jennifer L. Modliszewski, David L. Corcoran, José Jaime Martínez-Magaña, Janitza L. Montalvo-Ortiz, John D. Roache, Lynnette A. Averill, Stacey Young-McCaughan, Paulo R. Shiroma, Traumatic Stress Brain Research Group, David A. Lewis, Jill Glausier, Paul Holtzheimer, Matthew J. Friedman, Jing Zhang, Alan L. Peterson, Chadi G. Abdallah, Xinyu Zhang, Ke Xu, John H. Krystal, Ronald S. Duman, Hongyu Zhao, Douglas E. Williamson, Matthew J. Girgenti

## Abstract

Post-traumatic stress disorder is a mental disorder that may occur in the aftermath of severe psychological trauma. We examined 1,065,750 DNA methylation (DNAm) sites from 171 donors including neurotypicals, PTSD, and major depressive disorder cases across six areas implicated in the fear circuitry of the brain. We found significant differential methylation for PTSD near 195 genes and utilizing cross-region modeling, identified 6,641 candidate genes. Approximately 26% of differentially methylated CpGs were present near risk loci for PTSD. To identify potential therapeutic intersections for PTSD, we found significant methylation changes in the *MAD1L1, ELFN1*, and *WNT5A* genes in ketamine responders. Finally, to better understand the unique biology of PTSD, we analyzed matching methylation data for a cohort of MDD donors with no known history of trauma or PTSD. Our results implicate DNAm as an epigenetic mechanism underlying the molecular changes associated with the subcortical fear circuitry of the PTSD brain.

## INTRODUCTION

Post-traumatic stress disorder is a debilitating psychiatric disorder with an approximately 7% prevalence in the general population^1–3^. PTSD typically emerges following extremely stressful life events, such as direct threats to one’s life. Particularly when chronic, it is often comorbid with other psychiatric diagnoses including major depression^4^ and substance use disorder^5^. Because the diagnostic criteria for PTSD specify that it is a lasting clinical condition arising from discrete environmental exposures (trauma), epigenetic mechanisms are very likely to contribute substantially to its pathophysiology.

Methylation of DNA (specifically at cytosine nucleotides) is a critical, epigenetic regulator of genome architecture, gene expression, and cell function^6^. These processes are important for mammalian brain development^7^, aging^8^, disease^9^, and response to external stimuli such as stress^10^. Epigenetic changes evoked by stress are thus encoded onto the genome and can serve as a link between the genetic architecture and the response (i.e., gene expression). In this way, epigenetic changes are a path through which traumatic stress and other environmental exposures influence PTSD vulnerability and resilience and converge mechanistically with underlying risk for PTSD^11^. Elucidating this interplay is thus fundamental to advancing our understanding of the etiology and pathophysiology of PTSD. Previous studies of the PTSD methylome have predominantly examined peripheral tissues such as blood^12–15^. These findings have largely centered on changes in inflammatory/immune response and glucocorticoid signaling. However, because epigenetic changes are tissue-, region-, and cell-specific, peripheral studies are limited in their ability to inform the understanding of brain epigenetic regulation of gene expression.

In this study, we focused on subregions of the amygdala and the hippocampus because these are among the most well-understood components of the brain’s fear circuitry with known functional engagement in PTSD^16^. The amygdala is comprised of several nuclei with discrete functions including the basolateral nucleus (BLA), the central nucleus (CeA) and the medial nucleus (MeA). Neuroimaging and animal studies have found that the amygdala modulates the fear response^17–19^ and recent studies have demonstrated that individuals with PTSD exhibit greater amygdala activation relative to comparison subjects^20, 21^. Further, functional imaging studies have examined connectivity between the prefrontal cortex (PFC) and the amygdala and observed impaired inhibition in PTSD subjects^22^. The hippocampus is primarily involved in storing memory. Notably, gross hippocampal volume is decreased in PTSD patients compared to traumatized controls who did not develop PTSD^23^. We included three subregions of the hippocampus in our analysis: the dentate gyrus (DG), CA subfields (CA), and the subiculum (Sub).

We generated DNAm data from six postmortem primary brain regions from 171 individual donors using the targeted next-generation sequencing of bisulfite converted DNA (targeted methyl-seq). After rigorous quality control, this provided genomic coverage across 1,065,750 CpG sites representing 22,544 genes, in a much larger sample size (117 cases versus 54 controls) than previous studies, across two unique diagnostic groups (PTSD and major depressive disorder) compared to neurotypical controls in brain regions (subregions of the amygdala and hippocampus) not extensively examined in previous large DNA methylation cohorts. The goal of this effort was to comprehensively measure epigenetic responses (i.e., DNA methylation differences) across discrete brain regions with known involvement in PTSD pathophysiology. We have also included a non-PTSD psychiatric comparison group of individuals with (MDD) due to the high comorbidity between the two disorders^24^. We find differential DNAm signatures across all six regions including many non-overlapping, sex-specific differences. Many DNAm signals are present near genes regulating GABAergic transmission such as *ELFN1* which has previously been implicated in PTSD interneuron dysfunction^25^ and glucocorticoid signaling including *CRH1*. We also find that DNAm changes aggregate at genes previously implicated in genetic risk for PTSD and its associated clinical phenotypes including *KANSL1, MAD1L1,* and *CRHR1*^26^. Remarkably, we also identified significant changes in methylation of the *ELFN1, MAD1L1*, and *WNT5A* genes in response to the rapid-acting antidepressant ketamine in a cohort of PTSD patients. Taken together, this work represents a unique and powerful resource for exploring DNA methylation changes in human subcortical regions important for psychological stress pathology and provides critical neurobiological targets for future development of therapeutics.

## RESULTS

### Widespread DNAm differences in subcortical brain regions

We obtained high-quality DNAm data from 171 human postmortem individuals matched for sex, including donors diagnosed with PTSD, major depressive disorder (MDD), and neurotypical controls using targeted sequencing of bisulfite converted DNA (Targeted Methyl-Seq). We mapped the DNAm landscape of six postmortem brain regions: three amygdala nuclei: basolateral (BLA), central (CeA), and medial (MeA); and three hippocampal subregions: *Cornu Ammonis* (CA) subfields, dentate gyrus (DG), and subiculum (Sub) using a custom bioinformatic analysis pipeline (**Figure 1A** and **Github link**). We removed probes on the sex chromosomes, leaving 1,065,750 autosomal probes for analysis. Principal components (PC) analysis of PC1 versus PC2 found that sex had the greatest effect on methylation level variation (**Figure 1B**). Our cohort was well matched for sex (PTSD: 31 females, 30 males; MDD: 24 females, 33 males; and normal controls: 21 females, 32 males). We observed substantial variation in DNAm levels between all six regions. Notably, while we observed relatively distinct clusters for each subregion, amygdala (red, orange, and yellow) and hippocampal (blue, purple, and green) subregions tended to cluster together suggesting regional similarities in CpG methylation patterns (**Figure 1C**). Given the similarities in DNAm levels between subregions of the amygdala and hippocampus observed by PCA, we tested for unique DNAm signals for each subregion. For each subregion, we performed differential methylation analysis to compare each region to the other combined five regions. **Figure 1D** shows the heatmap of z-score transformed median methylation levels of the 1,200 CpG sites (top 200 most significant CpG sites for each of the six regions). Hierarchical clustering of these 1,200 sites reveals subregion similarities within each primary region. The top 200 significant CpGs are listed in **Supplementary Table 1**. Interestingly, few genes (24) overlapped between regions. The transcription factor *ZIC1* had shared CpGs across four regions including the CA, DG, subiculum and BLA which is likely consistent with its role in neurogenesis^27^. Taken together, these findings reveal region-specific differentially methylated CpGs (DMCs) within distinct genomic loci and biological pathways.

**Figure 1.**
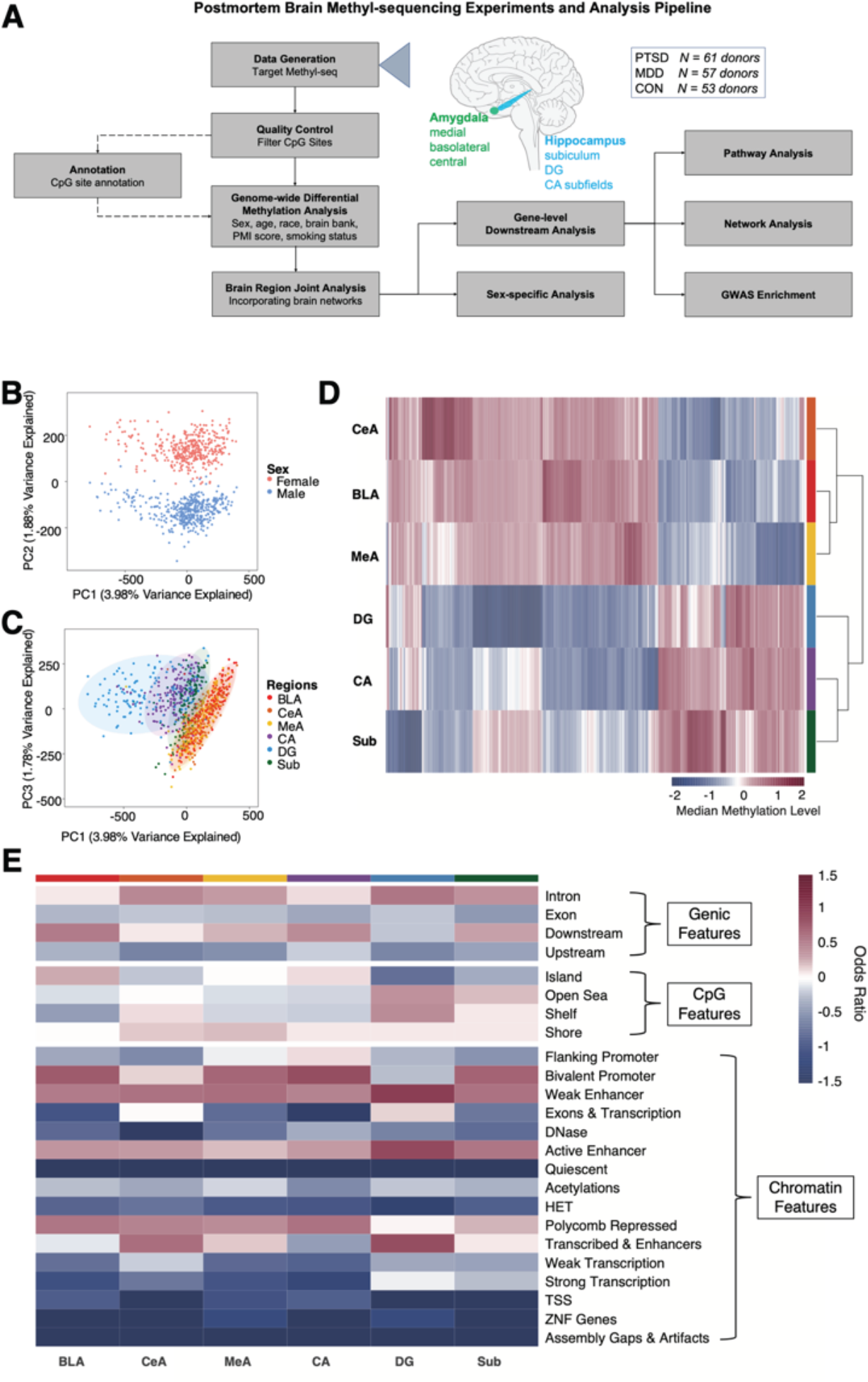
| DNA isolated from different brain regions display widespread differences in CpG methylation. **(A)** Bioinformatics analysis pipeline. **(B)** Individual scores from principal components analysis of all samples included in the study. PC1 and PC2 maximally separate subjects by sex. **(C)** PC1 and PC3 maximally separate subjects by brain regions. **(D)** heatmap of median methylation levels of the 1,200 CpG sites (top 200 most significant CpG sites for each region that distinguish each of the six brain regions from the five others); only samples with data from every brain region were included. Hierarchical clustering of these 1,200 sites reveals subregion similarities within each primary region. **(E)** Heatmap of *z*-score transformed odds ratios for genic, CpG and chromatin features showing the odds ratio of Bonferroni-corrected significant CpG sites within the feature versus excluded. Significant subcortical CpGs are more enriched in bivalent promoter, weak and active enhancer regions (*P* < 0.0001) and less enriched in TSS, ZNF genes, and gaps (*P* < 0.0001).

To better understand the regional differences in DNAm, we annotated each CpG site for its genic features, CpG features, and chromatin features. Odds ratios of Bonferroni-corrected significant CpG sites within each feature versus excluded were calculated (**Figure 1E**). Significant subcortical CpGs are more enriched in bivalent promoter regions (mean O.R.= 1.47, mean *P* < 0.0001), enhancer regions (weak (mean O.R. = 1.57, mean *P* < 0.0001) and active enhancers (mean O.R. = 1.39, mean *P* < 0.0001)) and less enriched in TSS (mean O.R. = 0.39, mean *P* < 0.0001), ZNF genes (mean O.R. = 0.32, mean *P* < 0.0001) and gaps (mean O.R.=0.11, mean *P* < 0.0001), consistent with previous studies examining CpG methylation patterns in other brain regions^28^.

### DNAm differences between PTSD subjects and neurotypical controls

We compared differences in CpG methylation levels for each region between our control and PTSD cohorts controlling for sex, age at death, ancestry, brain bank, PMI, and smoking status (**Extended Data Figure 1**). We used dispersion shrinkage for sequencing^29^ which was initially developed for the analysis of bisulfite sequencing data. In the combined sex analyses, we identified five FDR-significant DMCs (*P* < 0.05). We found one hypermethylated DMCs in the CA for the *SNAR-G2* gene and four hypomethylated DMCs for microRNA *miR54812* and *NDUFAF1, HORMAD2-AS1*, and *NXN* genes in the DG (**Figure 2A**). In females, we found six significant DMCs in the BLA, 10 in the CeA, 16 in the MeA, 19 in the CA, 86 in the DG, and 13 in the subiculum (**Figure 2B** shows the top ten most FDR significant sites; the full list is available in **Supplementary Table 2**). We also found non-overlapping male-specific gene-associated DMCs in the BLA (three genes), in the CeA (eight genes), in the MeA (11 genes), in the CA (11 genes), in the DG (15 genes), in the subiculum (16 genes) (**Figure 2C**). The most significant genes with DMCs for all regions and by sex-specificity are shown in **Table 1**.

**Figure 2.**
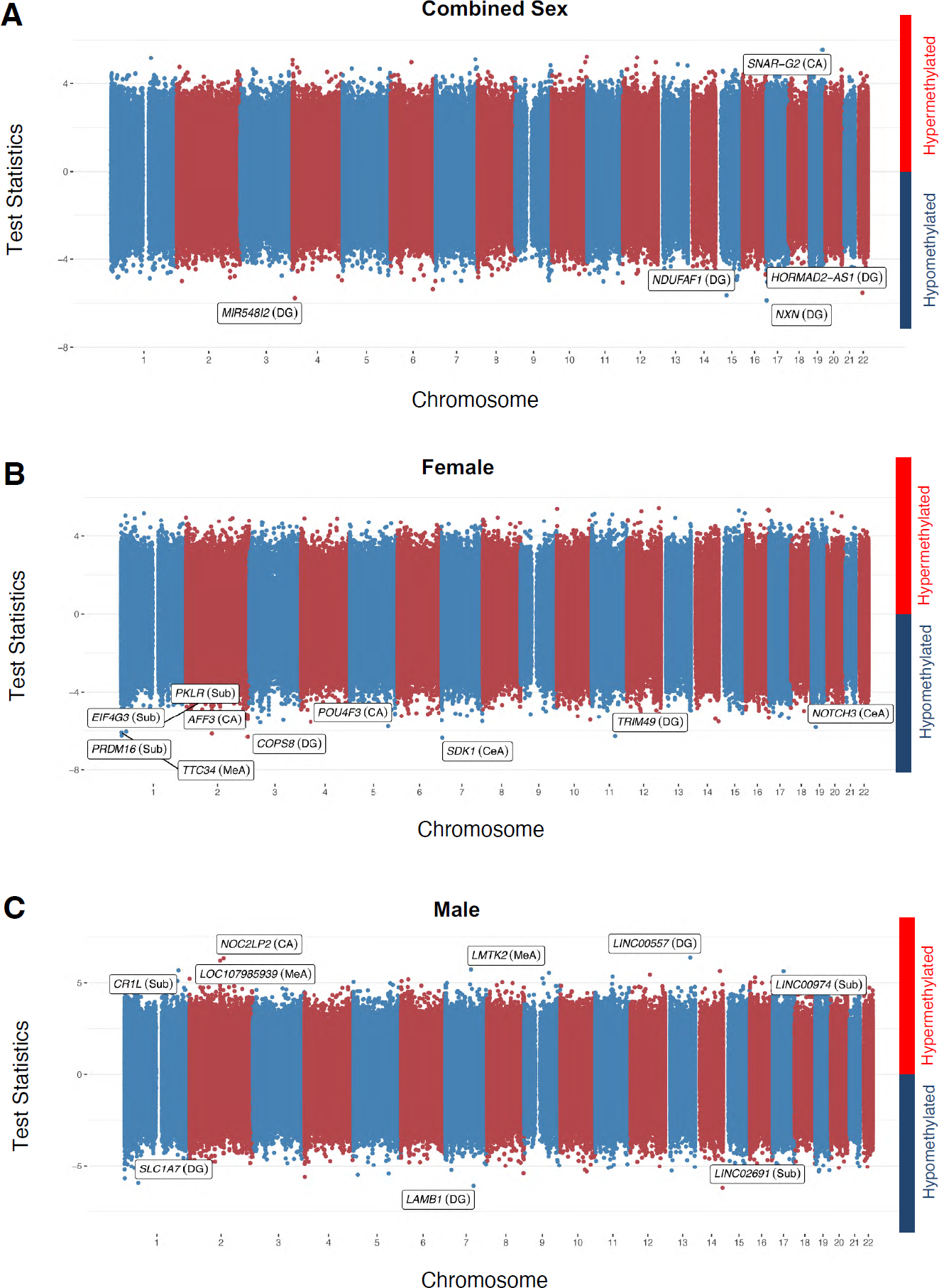
| Univariate analysis reveals regional and sex-specific differences in CpG methylation between PTSD cases and controls. **(A)** Two-sided Manhattan plot for PTSD case-control differential methylation using all samples included in the study. Differential methylation analysis was performed using a Bayesian hierarchical model, covarying for age, sex, ancestry, post-mortem interval, data source, and smoking status. The association test statistics for each variant tested is reported on the y-axis for hypermethylation (above) and hypomethylation (below). The top FDR significant markers across six regions are annotated. Labels indicate gene and regions for significant DNAm. **(B)** Two-sided Manhattan plot for PTSD case-control differential methylation using only female samples, covarying the same set of covariates excluding sex. **(C)** Manhattan plot for PTSD case-control differential methylation using only male samples, covarying the same set of covariates excluding sex. For female- and male-specific analysis, only top the ten most significant CpGs across six brain regions are annotated; the full list is available in **Supplementary Table 2.**

**Table 1.** The top 5 FDR significant DMCs in the univariate analysis for combined-sex, and two sex-specific analyses.

| Analysis | Chromosome | Position (bp) | Region | Closest Gene | Test Statistic | P |
| --- | --- | --- | --- | --- | --- | --- |
| Combined Sex | 19 | 799710 | DG | NXN | -5.8737 | 4.26E-09 |
|  | 6 | 9533078 | DG | MIR548I2 | -5.7705 | 7.90E-09 |
|  | 17 | 41403096 | DG | NDUFAF1 | -5.6408 | 1.69E-08 |
|  | 21 | 49029942 | CA | SNAR-G2 | 5.5576 | 2.73E-08 |
|  | 24 | 30080197 | DG | HORMAD2-AS1 | -5.5301 | 3.20E-08 |
| Female | 9 | 4221321 | CNA | SDK1 | -6.3581 | 2.04E-10 |
|  | 4 | 237157075 | DG | COPS8 | -6.3181 | 2.65E-10 |
|  | 13 | 89787631 | DG | TRIM49 | -6.2778 | 3.43E-10 |
|  | 3 | 3297053 | Sub | PRDM16 | -6.2539 | 4.00E-10 |
|  | 4 | 99593922 | CA | AFF3 | -6.1477 | 7.86E-10 |
| Male | 15 | 94968558 | DG | LINC00557 | 6.3734 | 1.85E-10 |
|  | 4 | 131443806 | CA | NOC2LP2 | 6.3396 | 2.30E-10 |
|  | 4 | 117859040 | MNA | LOC107985939 | 6.2142 | 5.16E-10 |
|  | 16 | 104317737 | Sub | LINC02691 | -6.2110 | 5.27E-10 |
|  | 9 | 107952197 | DG | LAMB1 | -6.1054 | 1.03E-09 |

### PTSD-associated JMCs identified by joint brain region analysis

To identify differentially methylated sites with common effects across multiple brain regions, we adopted a powerful cross-brain joint region analysis strategy. The existing methods for differential methylation analysis are mostly based on the analysis of a single marker (univariate analysis) that requires a significantly larger number of samples. In fact, we were only able to find a limited number of DMCs (and associated genes) between PTSD and controls in each brain region after the Benjamini-Hochberg correction (**Figure 2**). The Markov Random Field (MRF) model has been applied to both genome-wide association studies and bulk RNA-seq studies to model biological dependencies/networks in genomic and transcriptomic data^30–32^. In these previous studies, MRF modeling of cross-regional genomics was extensively simulated to limit the detection of false positives making it ideal for use in our postmortem methylation dataset. Therefore, we employed a statistical framework that incorporates network topology information to better classify each CpG site into different methylation patterns to identify shared epigenetically modified biological processes (**Figures 1C** and **1D**).

There are a total of 64 patterns for each sex-specific and combined sex-comparison (2^6^, for each region a site is either a DMC or not (two possibilities) across six subregions) (**Supplementary Table 3** and **Extended Data Figure 2**). Joint analysis revealed 17,481 joint-region methylated CpGs (JMCs) (across all six regions, mean of 2,914 JMCs per region) (out of a total of 1,065,750 CpGs or 0.27%) for all single region analyses in our combined-sex dataset. This corresponded to 1,761 nearest genes for BLA, 1,672 for CeA, 2,134 for MeA, 2,015 for CA, 1,687 for DG, and 1,859 for Sub (within 10 kb and nominally significant by univariate analysis) (**Supplementary Table 3**). Across all differential DNAm patterns, we identified 48,158 JMCs, of which 41.3% are significant DMCs in single subnuclei of the amygdala and 40.3% are significant in specific hippocampal subregions, indicating similar changes of DNAm across regions (**Extended Data Figure 2A**). Consistent with our univariate PCA analysis, most jointly shared DMCs fall within the combined amygdala (9%) or combined hippocampal comparisons (7.8%). Additionally, this pattern was observed in both the female-specific (**Extended Data Figure 2B**) and male-specific comparisons (**Extended Data Figure 2C**). Interestingly, we found 52 JMCs mapping to 36 genes common to all regions (**Supplementary Table 3**). Gene ontology analysis of each pattern was performed individually. We show the top 10 canonical pathways across the six individual subregion patterns in **Extended Data Figure 3** and found the greatest enrichment for pathways related to axon guidance signaling, transcription factor CREB signaling in neurons, GABA receptor signaling, and endocannabinoid signaling (*P* #x00FD; 0.01 for all gene sets).

We observed similar pathway enrichments in our sex-specific analyses (**Supplementary Table 4**) and in the multi-regional pattern analyses confirming our expectation that PTSD has similar associations with DNAm at many sites across different brain regions. The complete set of gene set enrichments for each region and joint-region comparison are in **Supplementary Table 4.**

**Figure 3.**
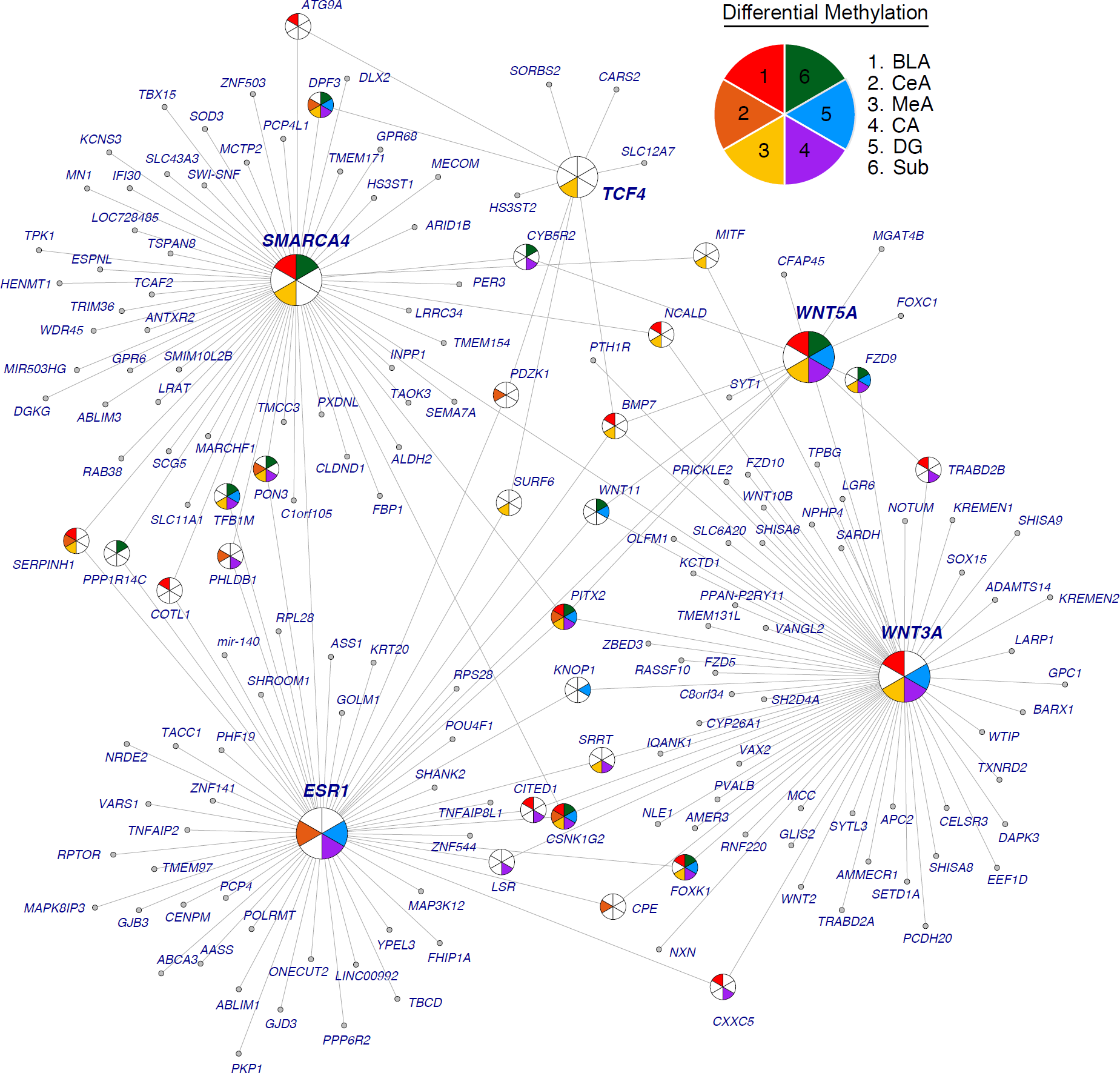
| Network analysis. Gene network analysis was performed using IPA for JMCs in each differential methylation pattern. The most significant gene networks (network score > 30) from the six single-regional analyses were obtained and merged by integrating the hub genes and their nearest neighbors in IPA. Pie-circles were used to indicate the differential methylation regions for genes that have more than two neighboring genes. The amygdala regions (BLA: red, CeA: orange, and MeA: yellow) are in warm colors and the three hippocampus regions (CA: purple, DG: blue, and Sub: green) are in cold colors. Genes *SMARCA4, ESR1, TCF4, WNT3A*, and *WNT5A* were hub genes with the greatest number of nearest neighbors.

### Gene network analysis reveals sex-specific and regional convergence of PTSD affected biological processes

In order to better understand how PTSD DMCs are affecting brain regions, we performed gene network analysis to identify possible convergent biological processes and interactions these DMC-associated genes are having. For each enrichment analysis, we extracted the most significant (network score > 30) gene networks for each single regional pattern and integrated hub genes and their nearest neighbors with the hub genes and neighbors of networks for multiple regions (**Figure 3**). Genes *SMARCA4, ESR1, TCF4, WNT3A*, and *WNT5A* were hub genes among the six regional patterns with the greatest number connected neighbors. We found significant DNA methylation changes in *SMARCA4* (*P* = 0.0007), *ESR1* (*P* = 0.0005), *TCF4* (*P* = 0.0004), *WNT3A* (*P* = 0.0003) *and WNT5A* (*P* = 0.0004). *SMARCA4* (60 connections) is a hub gene in the top networks for BLA, MeA and Sub regions. Several studies^33, 34^ have identified *SMARCA4* as a dysregulated gene in peripheral blood gene expression analyses of PTSD patients. Differences in *ESR1* (39 connections) were previously identified in a female-only cohort analyzing PTSD blood methylation levels^35^ and we identified a female-specific *ESR1* network that was not present in the male network analyses (**Extended Data Figures 4** and **5**), suggesting sex-specific biological network changes involving sex hormones in PTSD. *TCF4* (9 connections) is one of the most significant PTSD risk variants detected by GWAS^26, 36^ and is a significant hub gene in the MeA. We also found hub genes related to WNT signaling including *WNT3A* (66 connections) that was a hub in BLA, MeA, CA, and DG regions, and *WNT5A* (10 connections) which was a hub gene in five of the six regions. Previous studies have identified roles for WNT signaling in the impairment of fear extinction, excessive anxiety and in depression^37, 38^. Taken together, these data suggest convergent and highly coordinated disruption of gene networks in the fear circuitry of the PTSD brain that differs between males and females. The full list of gene network results is found in **Supplementary Table 5.**

**Figure 4.**
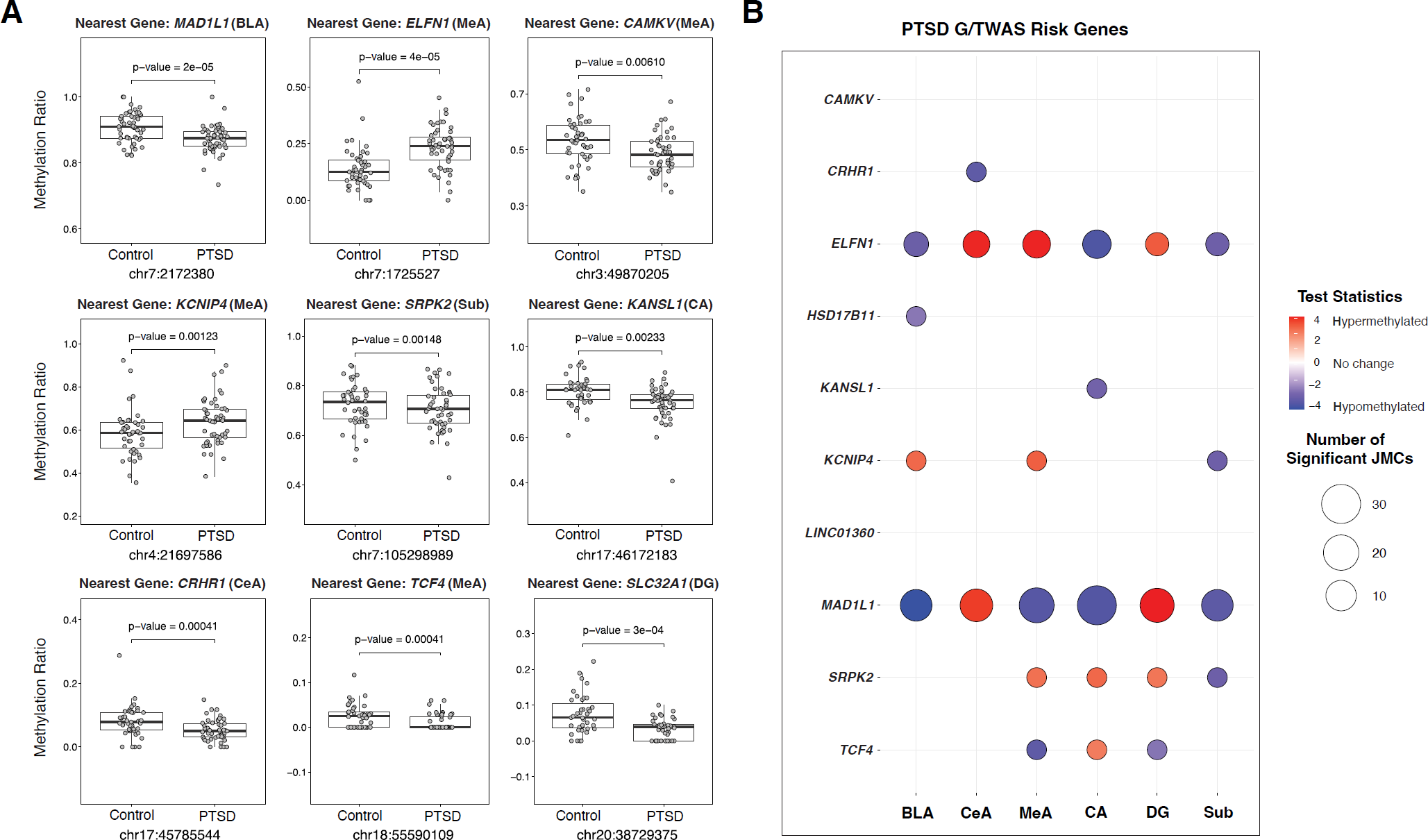
| Examples of DNAm changes for 10 GWAS-positive loci for PTSD. **(A)** Examples of group-specific DNA methylation ratios for ten PTSD risk genes. *MAD1L1, CAMKV, KCNIP4, SPRK2, KANSL1, CRHR1*, and *TCF4* are significant risk genes (identified by GWAS) for PTSD. *ELFN1* is a significant TWAS hit for PTSD, and *SLC32A1* is a previously identified transcriptomic key driver in PTSD. For each boxplot, y-axis dots show methylation levels at a specific CpG site. *P*-value corresponds to differential methylation. For box plots, center line is the median, limits are the IQR, and whiskers are 1.5X the IQR. **(B)** Methylation changes at nine genes identified by the largest PTSD GWAS using data from MVP were examined. Significant changes were found for *KCNIP4, HSD17B11, MAD1L1, SRPK2, KANSL1, CRHR1*, and *TCF4* for at least one brain subregion. Significant enrichment of DNAm changes were also found in the interneuron gene *ELFN1*, which was a PTSD genetically regulated genes identified by TWAS.

**Figure 5.**
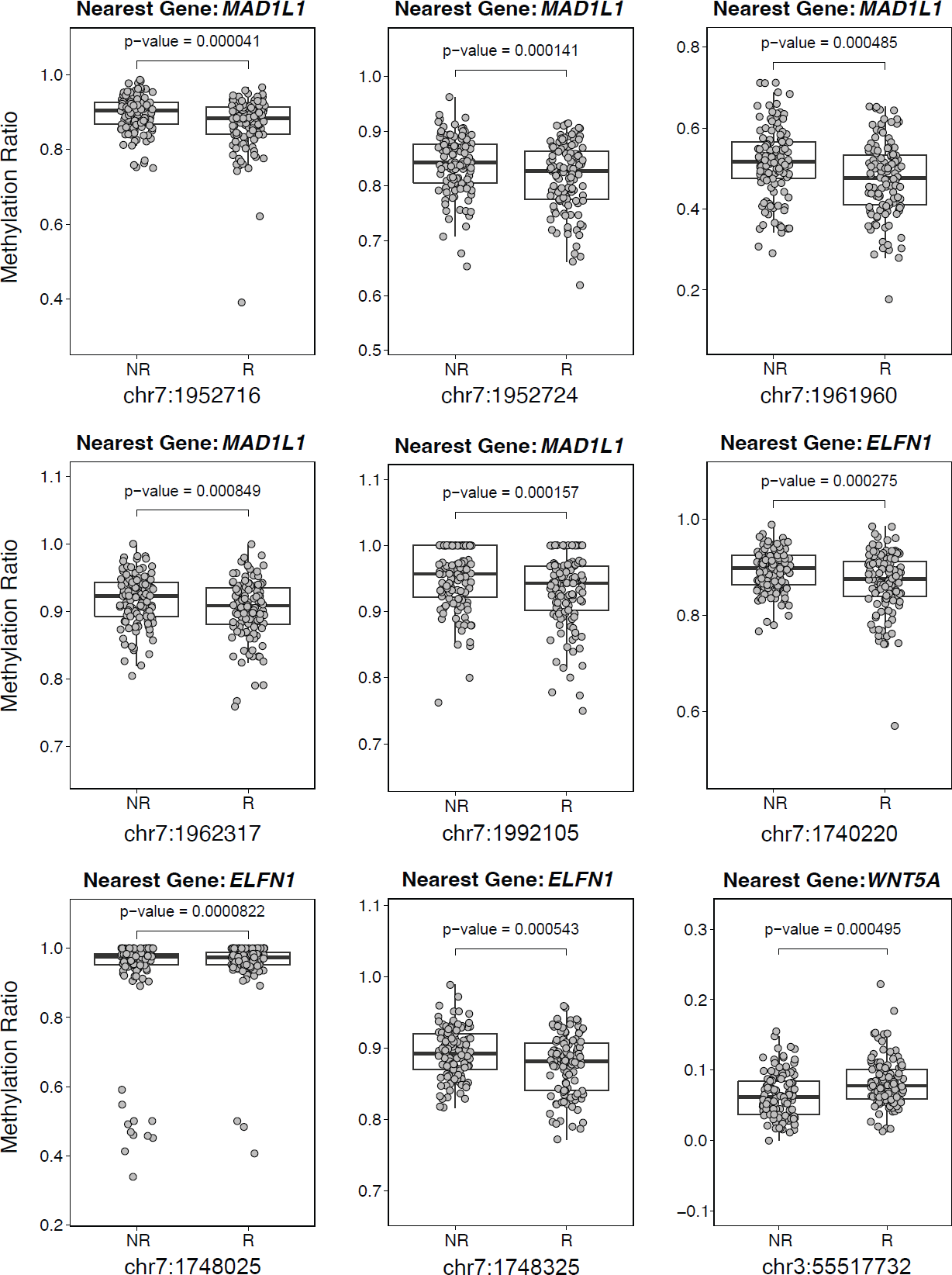
| Examples of ketamine altered DNA methylation ratios for three PTSD genes. Significant hypomethylation at five sites in the promoter of *MAD1L1* (*P* = 0.00004, 0.0002, 0.00014, 0.0005, 0.0008) in patients who responded to ketamine, three in the *ELFN1* promoter (*P* = 0.00008, 0.0003, 0.0005), and one in the *WNT5A* promoter (*P* = 0.0005). For each boxplot, y-axis dots show methylation levels at a specific CpG site. P value corresponds to differential methylation. For box plots, center line is the median, limits are the IQR, and whiskers are 1.5X the IQR.

### DNAm changes at PTSD risk loci

We next tested for enrichment between DMCs that display epigenetic differences associated with PTSD and genetic risk variants identified in genome-wide association study for PTSD^26, 36^. We downloaded and analyzed the entire NHGRI GWAS catalog^39^ for PTSD that included 20 studies and included 137 SNPs and 147 mapped genes. First, we performed GWAS enrichment to identify variant-gene-disease networks associated with our DMCs. We mapped our DMCs to SNPs within the MVP GWAS so that we could test association of our DMCs with disease variants in the DisGeNet database (See Methods)^40^. We identified a variant-gene-disease network centered on *MAD1L1*, its risk variants and its connections to Major Depression and Cardiovascular Disease. We examined whether the risk variants were specific for any diseases and found significant enrichment for mental disorders, digestive system diseases and cardiovascular disease (**Extended Data Figure 6**).

**Figure 6.**
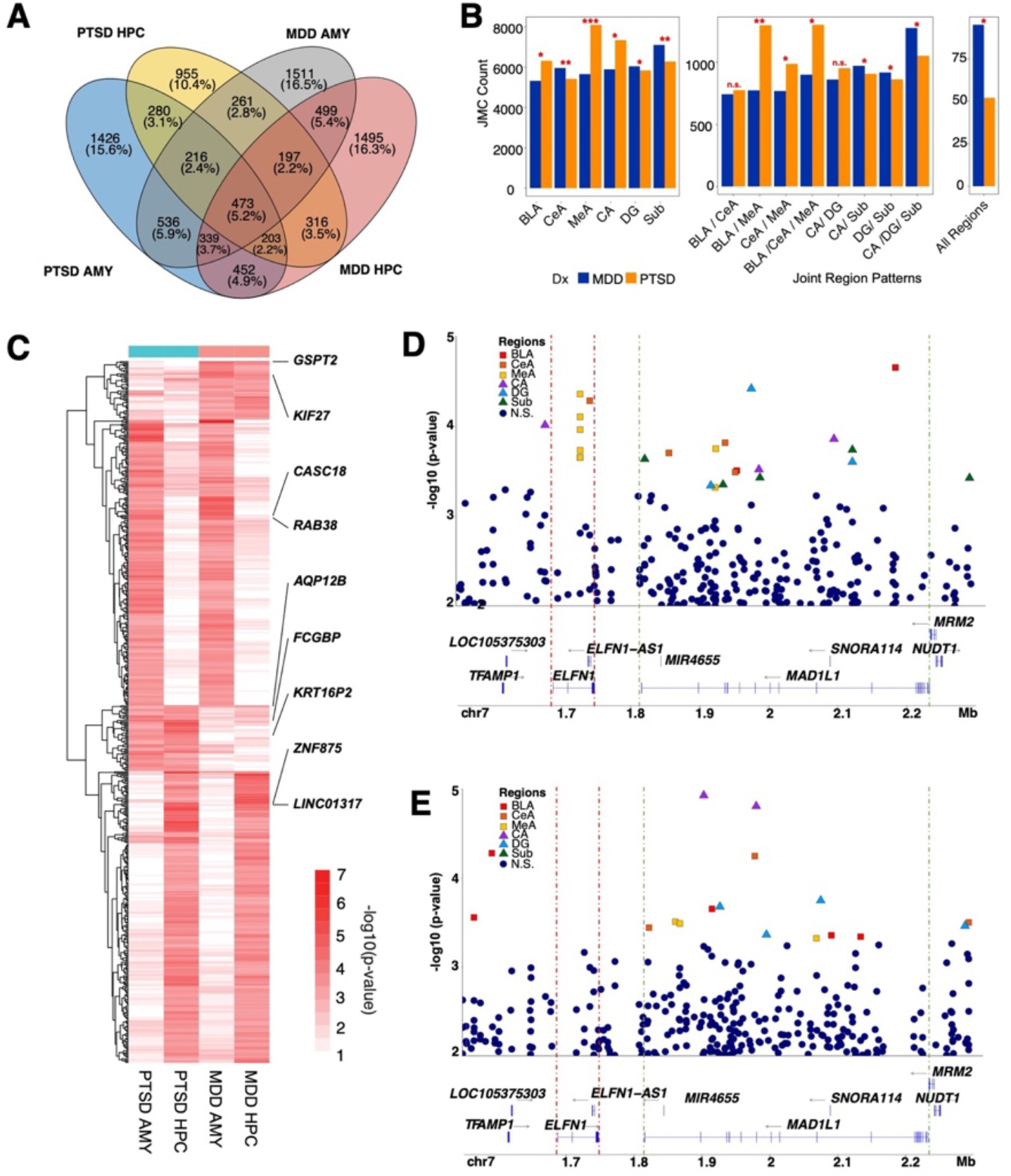
| Common and divergent epigenetic signatures for PTSD and MDD. **(A)** Venn diagram shows overlap of DMC-associated genes from the PTSD and MDD amygdala regions and hippocampus regions. The top 2,000 significant CpG sites from each region (BLA, CeA, MeA, CA, DG, Sub) were extracted from the univariate PTSD case-control differential methylation analysis and MDD case-control differential methylation analysis. We combined the sub-regional signals into amygdala and hippocampus primary regions. **(B)** –log10(p-values) are shown in the heatmap to show convergent and divergent genes for PTSD and MDD amygdala and hippocampus regions. We used all significant CpGs from the joint analysis as background CpGs. We used the mean p-value from three sub-regions to construct the unified p-value (amygdala: BLA, CNA, MNA; hippocampus: CA, DG, Sub). We plot CpG sites that have mean p-value less than 0.025. (**C**) Bar plot shows number of significant CpG sites in the joint analysis PTSD case-control analysis and MDD case-control analysis. \**P*<0.05, \*\**P*<1e-20, \*\*\**P*<1e-40. (**D**) Locus zoom-in plots on chromosome 7 for PTSD combined-sex analysis and (**E**) MDD combined-sex analysis. The significant hits in amygdala regions were plotted by subregion (red-BLA, orange-CeA, yellow-MeA, purple-CA, blue-DG, and green-subiculum. Non-significant hits are colored in dark blue color.

Second, we found that 11,470 CpG sites (of the 42,582 CpG sites (26.9%), *P* < 0.05) were significantly, differentially methylated within the LD regions spanning 137 PTSD genome-wide significant loci. We found significant differences in DNA methylation ratios for *MAD1L1* (*P* < 0.0001), *CAMKV* (*P* = 0.0061), *KCNIP4* (*P* = 0.0012), *KANSL1* (*P* = 0.0023), *CRHR1* (*P* = 0.0004), and *TCF4* (*P* = 0.0004) and suggestive changes in *HSD17B11* (*P* = 0.0114) and *SPRK2* (*P* = 0.0015) (**Figure 4A**). We specifically examined methylation changes at nine genes identified by the MVP PTSD GWAS^26^ and plotted their differential methylation for each subregion (**Figure 4B**). We found significant changes for *KCNIP4, HSD17B11, MAD1L1, SRPK2, KANSL1, CRHR1*, and *TCF4* for at least one brain subregion. Because we identified a large portion of DMCs near PTSD risk genes (26.9%), we reasoned it was likely that enrichment of DMCs maybe occurring near or within other genes implicated in PTSD pathophysiology. Therefore, we explored the possibility of enriched methylation differences within PTSD genetically regulated genes identified by TWAS^25^. We found significant enrichment of DNAm changes in the interneuron gene *ELFN1* (**Figure 4A**) (O.R. = 12.25; *P* < 0.0001)^25^.

Finally, we extended this analysis to include all of the key drivers identified in the cortical interneuron module (**Extended Data Figure 7**) and found significant differential methylation for several GABAergic markers including *SLC32A1, GAD1, GAD2, LHX6, PNOC, GABRA1, GABRA1, PVALB*, and *VIP*. These genes are significant transcriptomic molecular drivers and their epigenetic regulation in the current study indicates disruption of GABA signaling plays a key role in subcortical regions and maybe associated with genetic regulation of CpG methylation at *ELFN1*. Taken together, these findings suggest that DNA methylation level changes proximal to risk loci for PTSD may influence or possibly mediate the effect of genotype on clinical risk for many genome-wide significant loci.

### Ketamine alters blood DNAm signatures in PTSD patients

In order to link our findings with potential clinical significance, we explored the methylation patterns in peripheral blood of PTSD patients who responded to the rapid acting antidepressant ketamine^41^. Intravenous ketamine was administrated twice a week to 97 veterans and active-duty service members with PTSD for four weeks. PTSD severity was assessed by self-reported PTSD check list for DSM-V (PCL-5) prior to each ketamine infusion and 24h post first and last infusions. Responder status was determined by the slope of trajectory on the PCL-5 score. DNA methylation was measured at 24h post 1^st^ and 8^th^ ketamine infusions and 2-3 days post 4^th^ ketamine infusion. We found significant ketamine-induced methylation changes in the several PTSD associated genes identified in our study (**Figure 5**). We observed significant hypomethylation at five sites (*P* = 0.00004, 0.0002, 0.00014, 0.0005, 0.0008) in *MAD1L1*, three in *ELFN1* (*P* = 0.00008, 0.0003, 0.0005), and one hypermethylated in *WNT5A* (*P* = 0.0005). For complete details on the differential CpG methylation changes induced by ketamine see **Supplementary Table 6**. These single gene analyses suggest broader clinical implications for methylation changes in PTSD patients.

### Distinguishing DNAm associations of PTSD and MDD diagnoses

PTSD is highly co-morbid with major depressive disorder (MDD) with more than 50% of newly diagnosed cases of PTSD also experiencing depressive symptoms^24^. Therefore, we included analyses of an MDD cohort matched for age, sex, PMI, and with a similar history of drug use to disentangle the unique and divergent biological processes of both disorders. We analyzed this MDD cohort in the same manner as our PTSD cohort (**Figure 1A**) and corrected for the same covariates (age at death, sex, ancestry, brain bank, PMI score, and smoking status) to make relevant biological comparisons between the two diagnostic groups (**Extended Data Figure 8, Supplementary Tables 7 and 8**). Unsurprisingly, we found that between disorders, JMCs were most similar between the same regions than in any other regional or multi-regional comparison and unique for PTSD and MDD (**Extended Data Figures 9** and **10**). Therefore, we collapsed each subregion in our univariate analysis (**Figure 2** and **Extended Data Figure 8**) to directly compare the overlap in the DMCs and genes between the disorders. Few specific DMCs overlapped between PTSD and MDD with the most overlap observed in the amygdala (5.9%) and fewer overlapping in the hippocampus (3.5%) (**Figure 6A**).

Interestingly, we noted that the medial amygdala (MeA) had significantly more, albeit unique DMCs in PTSD donors (8,100 CpGs) compared to MDD donors (5,659 CpGs) (O.R. = 1.31, *P* < 0.0001). This finding also held in all joint amygdala subregion comparisons that included the MeA (BLA/MeA: O.R. = 1.53, *P* < 0.0001, CeA/MeA: O.R. = 1.17, *P* = 0.0012, BLA/CeA/MeA: O.R. = 1.32, *P* < 0.0001) (**Figure 6B**). By relaying olfactory information to hypothalamic nuclei responsible for reproduction and defense, the medial amygdala plays a significant role in emotional behaviors^42^. Interestingly, previous findings^43^ using a partially overlapping set of PTSD and neurotypical donors to the current study found that the MeA had far more significant differentially expressed genes (1,054 DEGs, FDR < 0.05) when compared to the number of DEGs found in the dorsolateral prefrontal cortex (209 DEGs), anterior cingulate cortex (43 DEGs) and the basolateral amygdala (62 DEGs) analyzed in the same study. Taken together, these findings suggest a unique role for the medial amygdala in the fear circuitry of the PTSD brain.

We next directly compared the genes associated with these DMCs (**Figure 6C**) to identify overlapping and convergent biological processes. We identified one gene that overlapped across all regions and both diagnostic cohorts: *AQP12B* (*P* = 0.0006). We averaged *P* values across all six regions for all nominally significant sites and found 239 genes that were convergent between the PTSD and MDD amygdala and in the hippocampus, 244 genes overlapped between MDD and PTSD. In diagnosis-specific regional comparisons, we found 46 DMC associated genes were significant across both amygdala and hippocampal regions in the MDD group and 55 were significant across both regions in the PTSD group (**Figure 6C**).

We found several significant DMCs in the *MAD1L1* locus, a major risk gene for both PTSD and MDD (**Figures 6D** and **6E**). *MAD1L1*’s nearest neighbor gene is *ELFN1* (approximately 68 kbp away) and both genes are in high linkage disequilibrium. Previous studies have identified significant changes in *ELFN1* gene expression in the dorsolateral prefrontal cortex of the PTSD brain and *ELFN1* was a significant TWAS hit in the same study^25^. Interestingly, we found multiple hits in the intronic and exonic regions of *ELFN1* that were specific to PTSD and not found in the MDD cohort where no significant DMCs were found. These findings reveal unique regulation of *ELFN1* in subcortical brain of individuals with PTSD and suggest alterations in GABAergic signaling that do not appear to be occurring in MDD.

## DISCUSSION

Here we present the most comprehensive DNA methylation study of the postmortem PTSD brain. By using targeted methyl-sequencing, we were able to leverage the specificity of a DNA microarray chip with the resolution of sequencing technology. We focused on brain regions that are commonly overlooked in epigenetic studies of postmortem tissue, specifically the amygdala and hippocampus. Our study exhaustively catalogued DNA methylation signals in specific subregions of the amygdala and the hippocampus, identifying common and distinct molecular profiles for each that provide a powerful and invaluable genomic resource for normal epigenetic states and variation in these brain regions. These data are made available via dbGaP.

Because each region of the brain has a unique cellular composition and regulatory landscape that contributes to neurobiological function, studying several brain regions simultaneously is essential to translating knowledge of epigenetic modifications into insight of molecular risk processes. We were able to cast a wide net to catch PTSD-related genomic alterations by examining the epigenomes of six brain subregions concurrently: three subnuclei of the amygdala and three subregions of the hippocampus. As much as we concentrated on “cross-region” differential methylation – differences present in many brain regions – we also found and analyzed a sizable number of sites that differed in methylation depending on the region. It is important to note that gene expression patterns might not be the same across amygdala and hippocampal brain areas, even if differential methylation is shared across them. We will likely gain further understanding of the molecular mechanisms underlying the diverse susceptibilities of specific brain areas underlying PTSD neuropathology by examining the distinctive transcriptional profiles of these brain regions. Nevertheless, a sizeable portion (26.9%) of the DNAm differences were enriched inside currently known PTSD genetic risk loci, suggesting that some of the DNAm differences revealed here are relevant to the etiology of PTSD and particular symptom clusters, such as reexperiencing. However, to more firmly identify which of these DNAm alterations are causative vs an epiphenomenon in the brains of PTSD patients, more mechanistic investigations to better understand the co-occurrence of DNAm and expression changes are required.

Univariate analysis of our dataset identified a handful of genes with significant DMCs, a caveat noted in other postmortem methylation studies^44^. Previous work from our group and others have successfully applied “Joint Analysis” to transcriptomic and other high dimensional genomic data to identify significant biological signals where power maybe lacking^30, 31, 45, 46^. Therefore, we employed a Markov random field model joint analysis to take advantage of the regional colocalization of sub nuclei of the amygdala and subregions of the hippocampus and integrate univariate summary statistics from differential methylation analysis with topology information from these regions in order to improve our power in detecting biologically significant DMCs. Consequently, in our combined-sex analysis we were able to identify 6,641 genes with significant DMCs across six brain regions in PTSD and 6,462 in MDD, as opposed to five DMC-associated genes using simple univariate analysis. We anticipate expanded application of this strategy in future genomic studies.

To better understand the biological processes underlying the differential methylation signals we identified, we performed gene network analysis using Ingenuity Pathway Analysis (IPA) from Qiagen^47^. IPA curates a large database of empirically derived biological interactions providing a powerful tool for identifying how genes (in this case genes that differentially methylated in PTSD) are coordinated in regulating biological process and identifying hub genes with significantly large numbers of biological connections to other genes. Our gene network analysis identified several hub genes including *TCF4* (Transcription Factor 4). *TCF4* is a genetic risk gene for PTSD^36^ that is involved in transcriptional regulation during neurodevelopment and has long been known as a risk gene for schizophrenia^48^. We observed enrichment for DMCs associated with risk loci for PTSD (26.9%) suggesting a genetic regulation of CpG methylation in diseased tissue. We observed significant DMCs within the *MAD1L1* gene that is a significant risk gene for PTSD^26^, MDD^49^, Schizophrenia and Bipolar Disorder^50^ pointing to a convergent role across neuropsychiatric disorders. However, our results also point to significant changes in methylation patterns in *MAD1L1*’s neighboring gene *ELFN1* that appears to be PTSD-specific (**Figures 6A** and **6B**). We have previously reported *ELFN1* gene expression changes in the frontal cortex of PTSD and MDD patients^25^. To date, no evidence has emerged implicating *ELFN1* involvement in these disorders stemming from subcortical gene expression dysregulation. The lack of significantly methylated CpGs in *ELFN1* in our MDD comparison group strongly suggests a unique role in the amygdala and hippocampus of PTSD subjects. Further studies of GABAergic transmission disruption are thus warranted.

Many studies have suggested that DNA methylation can be altered by antidepressants and promote symptom remission^51^. We therefore reasoned that epigenetic modification of PTSD genes with DMCs could be clinically relevant. Therefore, we explored changes in DNA methylation in the peripheral blood of PTSD patients treated with the rapid acting antidepressant ketamine (both responders and non-responders), with the aim of understanding the potential contribution antidepressants might play in the epigenome of PTSD. PTSD patient responders to ketamine had significant decreases in methylation in three PTSD hub genes (*ELFN1, MAD1L1*, and *WNT5A*) compared to those who did not respond to ketamine. Ketamine treatment response is based on DNA methylation changes across three time points, long after the drug was fully metabolized, suggesting longer term epigenetic changes. Previous studies have identified hypomethylation of the *MAD1L1* gene associated with PTSD among post-combat male military cohorts^52^. Further investigation into the relationship between ketamine, DNA methylation, and PTSD is needed to identify genome-wide epigenetic changes associated with rapid acting antidepressant administration.

In summary, we present results from the largest DNA methylation study of human brain subcortical regions to date. These results support the role of DNAm as a critical epigenetic modulator of gene expression in PTSD etiology. We observed sex-specific changes in the methylome males and females with PTSD. DNAm changes aggregated near PTSD risk loci and PTSD risk genes were major hub genes in gene networks suggesting a genetic link to downstream molecular functions. Further, we identified convergent and divergent epigenetic regulatory mechanisms between PTSD and MDD highlighting common pathways and genetic risk links. More generally, we illustrate the value of integrating epigenetic data across multiple brain regions to study complex psychiatric disorders.

## ONLINE METHODS

### Donors and Tissue

Human postmortem brain tissue samples were obtained from the National Center for PTSD Brain Bank (with consent of next of kin) and the University of Pittsburgh Tissue Donation Program. Individuals were a mix of European– and African–American descent. Males and females were group-matched for age and PMI. Sociodemographic and clinical details are listed in **Extended Data Figure 1** and include the cause and manner of death and comorbidities such as tobacco use and substances of abuse. Inclusion criteria for PTSD, MDD and control cases were as follows: PMI < 48h, age range >18 to < 85 years. A total of 171 individuals (61 PTSD: 31 females, 30 males; 57 MDD: 24 females, 33 males; and 53 healthy controls: 21 females, 32 males) were used in this study. The brain tissue was fresh frozen and samples from the basolateral nucleus (20 mg), basomedial nucleus (10mg), and central nucleus (10 mg) of the amygdala, the subiculum (20 mg), and the dentate gyrus (10 mg) and combined CA regions of the hippocampus (10 mg)^53^ were collected from each postmortem sample.

Psychiatric history and demographic information were obtained by psychological autopsies performed postmortem as well as a review of medical records and toxicology reports. Trained clinicians conducted structured interviews with informants (usually the next of kin) with knowledge of the deceased individuals. To avoid systematic biases, PTSD, MDD and control cases from each source were characterized by the same psychological methods. Consensus DSM-IV diagnoses for each subject were made by trained clinicians who did not conduct the psychological autopsies. Of the 61 individuals in the PTSD cohort, 82% (28 cases) were also comorbid for depression.

### Targeted Methylation Enrichment

Targeted next-generation sequencing of bisulfite converted DNA (targeted Methyl-seq) was achieved using the Roche KAPA HyperPrep Kits in combination with the SeqCap Epi CpGiant Probes. A panel of more than 2.1 million long oligonucleotide DNA probes (total capture size of 80.5 Mb) were used to interrogate >5.5 million methylation sites per sample at single-nucleotide resolution.

For each sample, DNA concentration was measured using a ThermoFisher Scientific Qubit Fluorometer and 250ng was subsequently fragmented using a Covaris E210 Focused-ultrasonicator to generate ∼300bp DNA fragments. Fragmented DNA samples were then converted into Illumina libraries using the KAPA HyperPrep Kit. In brief, DNA fragments were end repaired, A-tailed and ligated to barcoded Illumina adapters. Each library was then bisulfite converted using the EZ DNA Methylation-Lightning Kit by Zymo Research and amplified by polymerase chain reaction (PCR).

The bisulfite converted libraries were then hybridized with the SeqCap Epi CpGiant Probes in order to enrich the libraries for the > 5.5 million methylation sites across the genome. Enriched libraires were subsequently amplified by PCR before being pooled for sequencing on an Illumina NovaSeq 6000 Sequencing System. Three pools of 332 samples each were prepared. Each pool was sequenced on a NovaSeq 6000 S4 flow cell at 150bp paired-end, generating about 9Gb of data per sample.

### Blood Methylation Sample Description

Blood methylation analysis was a sub-study from the ketamine treatment on PTSD cohort^41^. DNA samples for DNA methylation in blood were from a double-blind, randomized, controlled trial study that was designed to examine ketamine treatment response among veterans and active-duty service members with PTSD. The participants (N = 158) were randomly administrated intravenous infusion of either placebo or ketamine for eight sessions. Only participants treated with ketamine (N = 97) were included in this analysis. Self-reported PTSD Checklist for DSM-5 (PCL-5) was used to assess PTSD symptoms. A slope from four weeks PCL-5 assessments were estimated using linear mixed regression model. Median of slope distribution was -1.98. Ketamine responder was defined as slope < -1.98 and non-responders was defined as slope > -1.98. DNA was extracted from whole blood. A total of 242 samples from three timepoints, day 1, 14, and 25 post first administration were used to assess ketamine response. Methylation was profiled by Agilent Sure SelectX Methyl-seq library preparation. Data processing and quality control followed our previous reported method^54^. Methylation at the sites near or in three genes, *MAD1L1, ELFN1*, and *WNT5A*, were extracted for association analysis for ketamine treatment response using linear mixed regression model, where timepoint was a random effect and covaried with age, sex, and self-reported ancestry.

### Bioinformatics

#### Data Processing & Quality Control

Raw data was processed using the fastp^55^ algorithm to trim low-quality bases and adapter sequences. Read-pairs that were still at least 200nt in length were mapped to the hg38 version of the human genome using the BSMAP algorithm^56^. Amplification artifacts were removed using the *MarkDuplicates* application from the Picard toolkit. The *methRatio* application from BSMAP was then used to call depth and percent methylation at each CpG loci in the human genome. CpG sites with a depth lower than 10x or greater than 125x within any sample were considered missing in that sample. Sites that are not present in at least 90% of the cohort were excluded from subsequent analysis. The median coverage of all sites for each sample were then scaled to the maximum median coverage of all samples. CpG sites were further excluded if they matched a common SNP as defined by dbSNP^57^ build 155. Finally, CpG sites that were considered invariant were excluded from analysis. Invariance was defined by CpG sites whose difference in the 10^th^ percentile from the 90^th^ percentile was less than a 5% change in methylation across the whole cohort CpG sites were annotated with the nearest gene based on their genomic coordinates in the hg38 version of the human genome (Ensembl^58^ version 96).

#### Assessment of Technical and Biological Variation

To assess for the presence of outliers, explore group structure and identify potential covariates, PCA was performed using the *pcaMethods*^59^ Bioconductor^60^ package in R. Associations between demographic and technical covariates and the first five principal components were tested utilizing an ANOVA for categorical covariates and a general linear model for numeric covariates. Association tests between group status (Control/PTSD/MDD) and covariates were conducted utilizing a general linear model for numeric covariates and Fisher’s Exact Test for categorical covariates. All *p*-values were adjusted for multiple testing via the Benjamini-Hochberg procedure (also referred to as FDR).

Each CpG site was annotated for its genic features, CpG features, and chromatin features. For genic features, we used the most recent human genome sequence datasets generated by UCSC (hg38) that included five genic features including intron, exon, promoter, upstream, and downstream. For CpG features, we also used the most updated hg38 reference database^61^ that included the following features: island, shore, shelf, and open sea. For chromatin features, we used the most recent universal chromatin state annotation of the genome, that uses a ChromHMM model and unifies over 1000 datasets^62^ that are divided into 16 chromatin features including weak enhancers, enhancers, transcribed & enhancer, weak transcription, transcription, exon & transcription, bivalent promoter, flanking promoter, quiescent, HET, polycomb repressed, acetylation, transcription start site (TSS), DNase, ZNF genes, assembly gaps & artifacts. Odds ratios of Bonferroni-corrected significant CpG sites within each feature versus excluded were calculated.

#### Differentially Methylated CpG Sites

To identify potentially differentially methylated CpG sites (DMCs) between control and PTSD and control and MDD individuals, a beta binomial linear regression model with an arc-sine link function was performed on each CpG site individually utilizing the *DSS*^29^ Bioconductor package in R. Age, sex, post-mortem interval (PMI), smoking status, ancestry, and tissue source were included as co-factors in the model. The models were run separately for each brain region and also for males and females jointly and individually (sex was excluded as a co-factor in the sex-specific models). To identify differentially methylated CpG sites across brain regions, six different sets of beta-binomial linear regression models were run. For each brain region, the region of interest was included as one factor level and the five other brain regions were considered to be the other factor level. SubjectID was included as a covariate in the model and only those subjects with data from all six brain regions were included.

#### Cross-region, Joint Differential Methylation Analysis

We utilized the nearest neighbor Gibbs measure to model the topology of brain network, which subsequently defines a Markov Random Field (MRF) model. We propose and implement an efficient algorithm that uses an Empirical Bayes method to maximize the pseudo-conditional likelihood for computational efficiency. Under mild regularity conditions, Besag^63^ showed that the maximum pseudo likelihood estimator is consistent and more efficient.

Summary statistics (*p*-values) from univariate analysis are used for input. The dimensions are 1,073,960 CpG sites by six brain regions. We performed filtering to remove CpG sites that have at least one missing *p*-value. 43,167 (about 4%) CpG sites were removed. Joint analysis was performed using a Markov Random Field model incorporating summary statistics from differential methylation analysis and topology information among brain regions. The CpG sites were classified into 64 methylation patterns.

For each CpG site, we assume that there is a known network between six brain regions. The network is represented by an undirected graph *G* = (*V*, ℰ) where *V* = {1, …, *B*} is a set of brain regions, here *B* = 6; ℰ = {< *i, j* >: *i* and *j* are directly connected} denotes the set of all edges. For brain region *i* in *V*, let *N_i_* = {*j*: < *i, j* > ∈ ℰ} denote the set of its neighbors, and *d_i_* = *N_i_*| denote the number of its neighbors. The true DM status for each CpG site is unknown, and we let *X_i_* denote the true association status where *X_i_* = +1 if brain region *i* is differentially methylated in PTSD, and *X_i_* = −1 if brain region *i* is NOT differentially methylated in PTSD. Let *X* = (*X*_1_, *X*_2_, …, *X*_6_) denote the node of each brain region. There is a total of 2^6^ = 64 unique configurations of the network. The objective is to infer the value of *X_i_* based on the brain network topology and the observed DM evidence. We adapt the statistical approach proposed by Chen et al.^64^ to improve statistical power to detect differential methylation patterns across brain regions. They used connectivity information among genes in a pathway to increase statistical power in GWAS analysis. We utilize the nearest neighbor Gibbs measure to model that CpG sites in connected brain regions tend to have similar DM status,

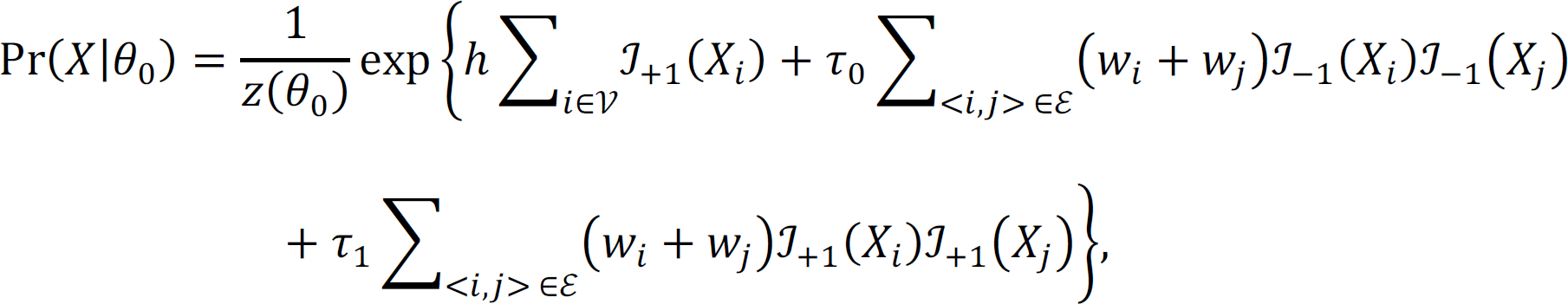

where θ_0_ = (ℎ, τ_0_, τ_1_) are hyperparameters; ℐ is an indicator function, i.e., when *X_i_* = +1, 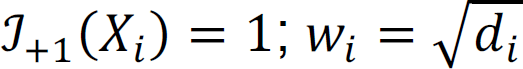; and *z*(θ_0_) is a normalizing function that is the sum over all 2*^n^* possible configurations. The observed statistical evidence for DM is summarized by p-values obtained from the univariate analysis when we treat each brain region separately. In addition, we consider the transformation that 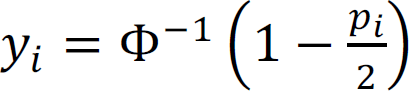, where Φ is the cumulative distribution function of a standard normal distribution. Under the null hypothesis of no association, we assume that *f*_0_(*y_i_*) ∼ *N*(0, 1). In contrast, where there is association, we assume that 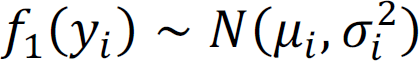. Here μ*_i_* and 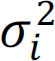 are unknown parameters and we put conjugate priors for μ*_i_* and 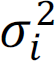,

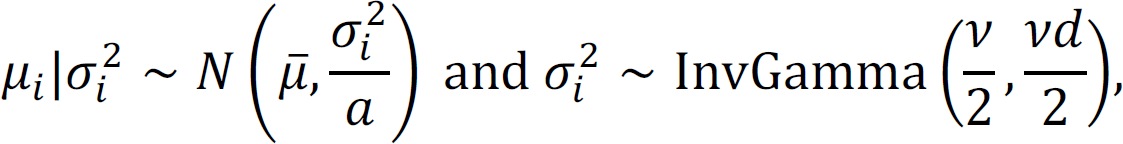

where we denote 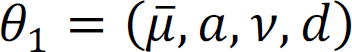. The marginal density of *y_i_* is

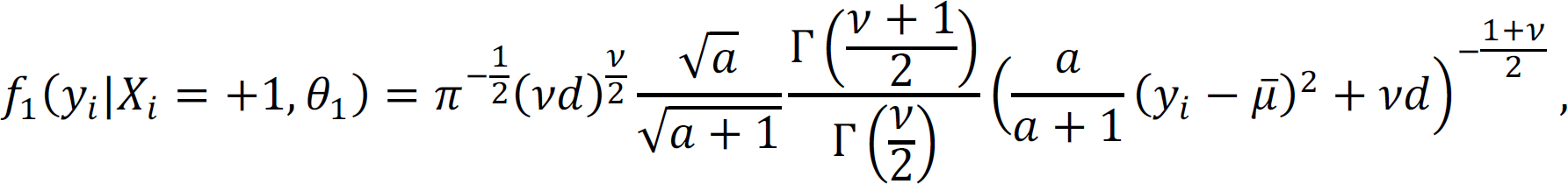

and the corresponding joint marginal density of *y* is

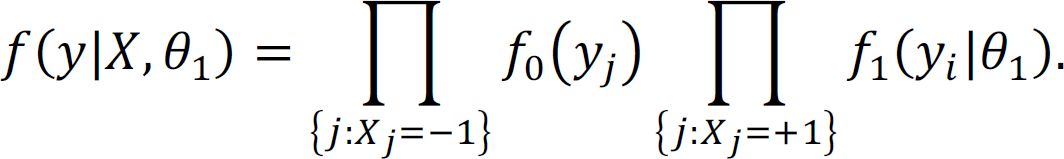

Therefore, according to Bayes’ theorem, the posterior distribution of *X* given *y* is proportional to

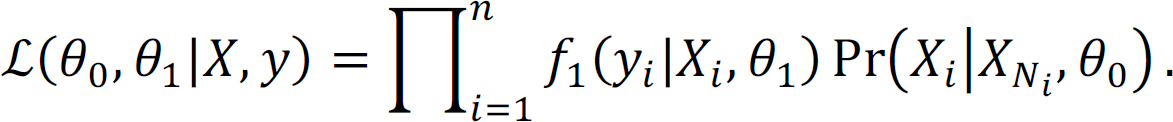

The hyperparameters can be estimated using the Empirical Bayes method by maximizing the pseudo conditional likelihood for computational efficiency,

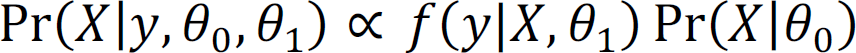

Under mild regularity conditions, Besag showed that the maximum pseudo likelihood estimator is consistent and more efficient. We estimate θ_“_ as follows: 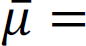 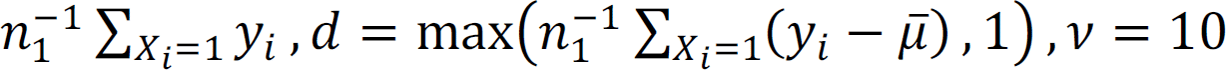 and 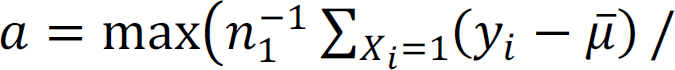 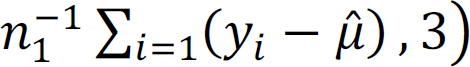. Meanwhile, θ_%_ can be estimated by maximizing 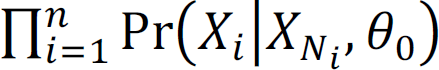. After we obtain hyperparameter estimates 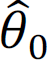 and 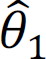, *X* can be sequentially updated by fixing 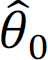 and 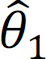. We use the following algorithm to obtain the final DM configuration that maximizes the pseudo conditional likelihood:

1. Randomly set an initial configuration *X*^(0)^;
2. In the *k*;^<^ iteration, obtain 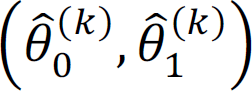 using *X*^(*k*–1)^;
3. Given 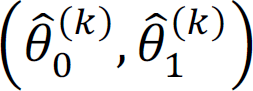, sequentially update the labels of *X*^(=)^ that maximize the pseudo conditional likelihood;
4. Repeat steps 2 and 3 until convergence;
5. Repeat steps 1 - 4 with different initial configurations;
6. Obtain the joint methylation pattern.

#### Gene Set Enrichment Analysis

To identify pathways and processes governing differences in Control vs. PTSD and Control vs. MDD, pathway analysis and GO enrichment analyses were performed. A p-value cut-off of 1e-4 was set as a threshold for inclusion of CpG sites in enrichment analyses for each model set run; this cutoff allowed for the inclusion of ∼100-500 unique genes in each enrichment analysis. Canonical Pathways Analysis was run in Ingenuity Pathway Analysis (IPA, QIAGEN); due to some genes being represented by more than one CpG site, the analysis was conducted using only p-value. In this case, only the hypergeometric test for pathway membership overlap is conducted and there is no independent test of directionality or corresponding z-score. DMCs in 64 differential methylation patterns (**Extended Data Figure 2**) were annotated to genes and IPA was performed on the mapped genes based on their relative genomic locations. Genes not mapped to the IPA database were excluded in our pathway analysis.

#### Gene network analysis

We also performed gene network analysis using IPA tools for six single-regional differential methylation patterns. Molecules are represented as nodes, and the biological relationship between two nodes is represented as an edge. All edges are supported by at least one reference that is in the IPA human database. The most significant network for each pattern was obtained and then merged using IPA. Pie-circles are used to indicate the differential methylation regions for genes that have more than two neighboring genes.

#### GWAS Enrichment of PTSD CpGs

We analyzed the published 137 genomic loci for PTSD risk corresponding to 147 associated genes in the NHGRI GWAS catalog. We calculated the proportion of DMCs that fall into mapped linkage disequilibrium (LD) regions to determine whether CpGs within PTSD risk loci differed in their DNAm effects. Specifically, we compared median differential methylation effect sizes between PTSD and normal control donors for each brain region.

#### Sex-specific Analysis

Sex is an important confounder in psychiatric disorder studies, especially for PTSD. We include sex as a confounding variable in the combined-sex differential methylation analysis and subsequent downstream analyses. We also performed sex-specific comparisons. We used only female samples and male samples to perform female-specific and male-specific differential methylation analysis, respectively. We then perform downstream analyses including enrichment analysis and gene network analysis using these sex-specific results.

## Data Availability

All data produced in the present study are available upon reasonable request to the authors

## Extended Data Figures

**Extended Data Figure 1.**
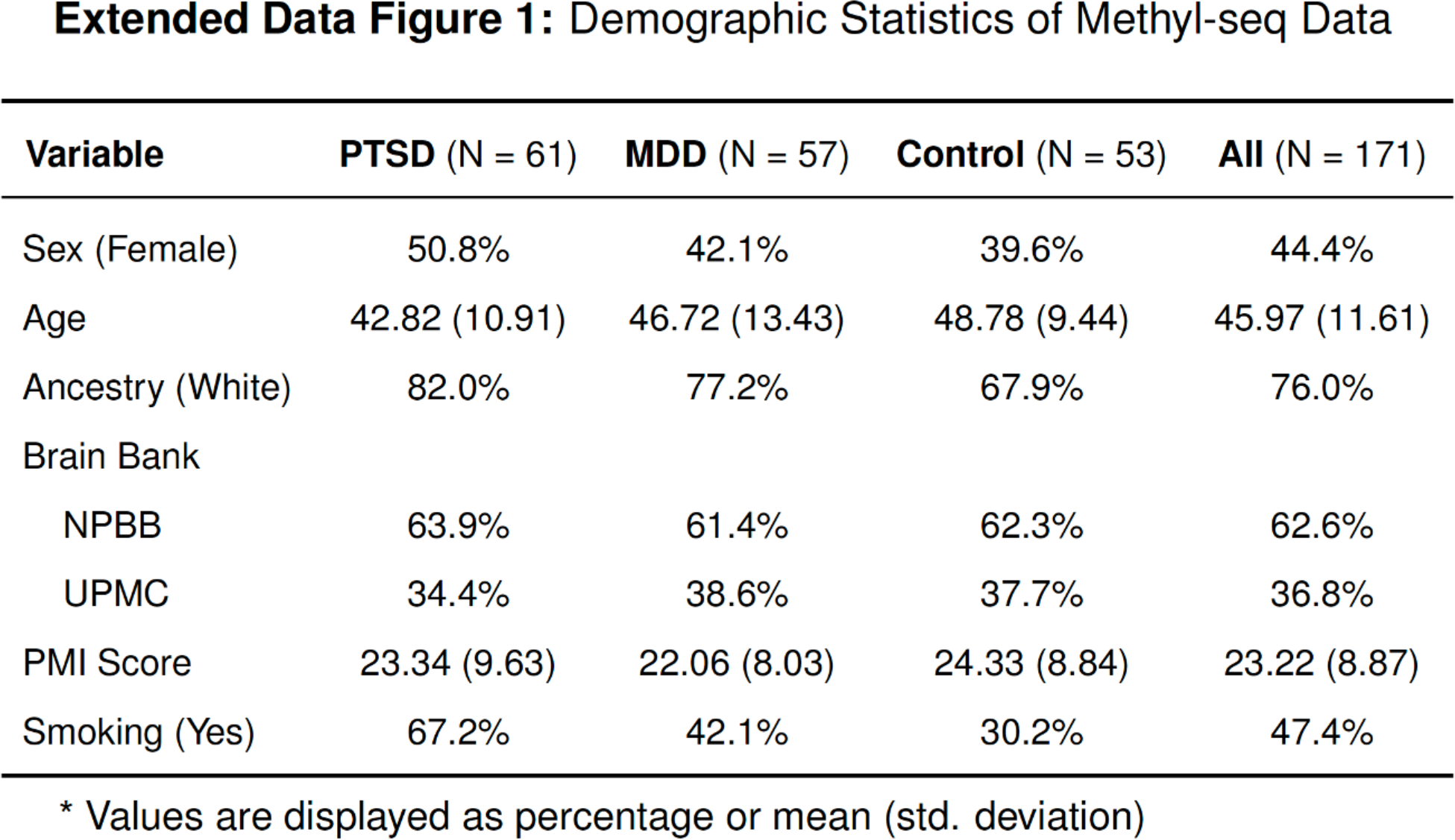
Postmortem Donor Demographics. Demographics in the study stratified by PTSD, MDD, and normal control groups. Sex, age at death, ancestry, brain bank, PMI score, smoking status were recorded.

**Extended Data Figure 2.**
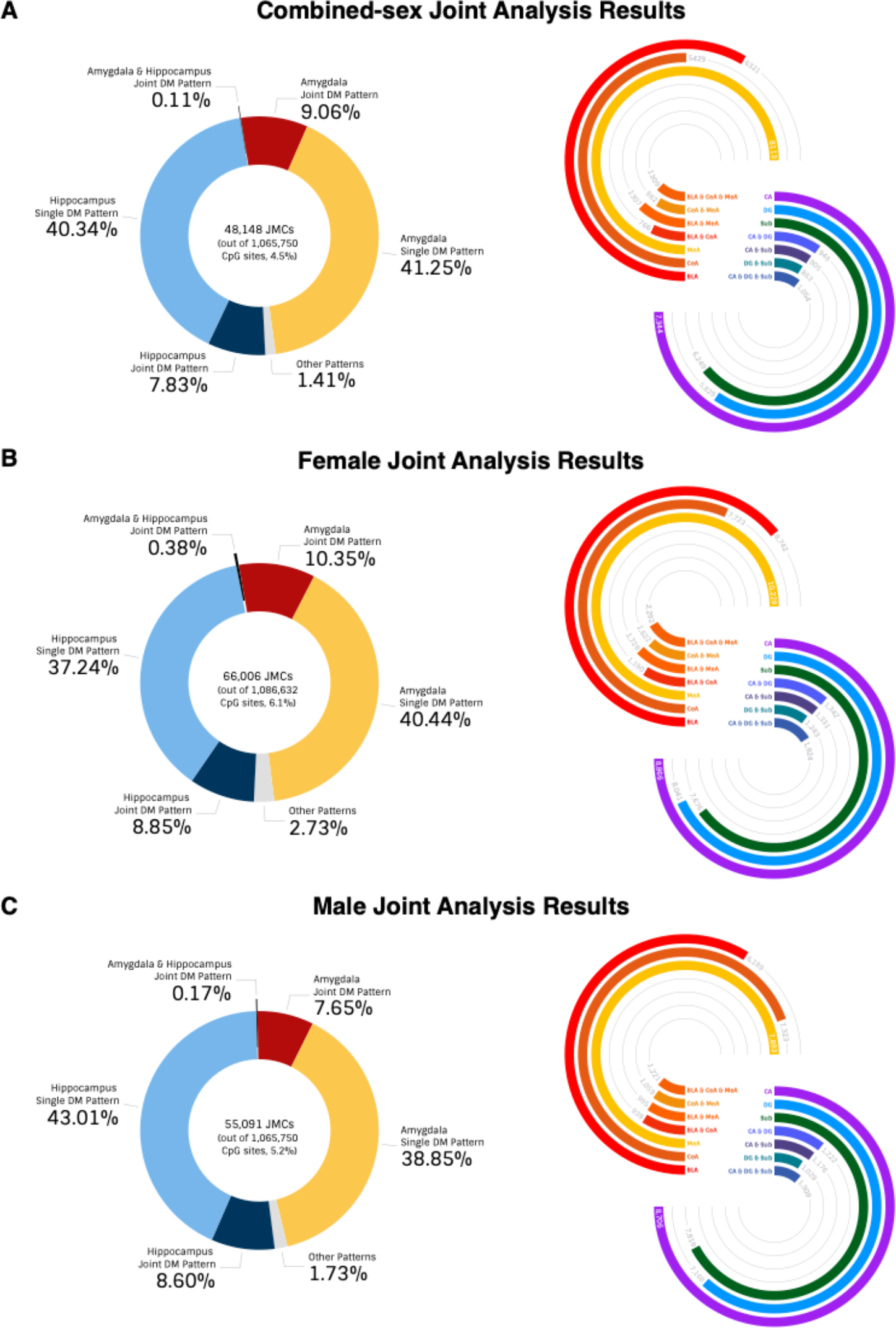
Joint analysis results for PTSD case-control combined-sex analysis. **(A)** Joint analysis was performed using a Markov Random Field model incorporating summary statistics from genome-wide differential methylation analysis and topology information among brain regions. The CpG sites are classified into 64 methylation patterns. The complete list of differential methylation patterns for each CpG site can be found in **Supplementary Table 3. (B)** Joint analysis results for PTSD case-control female analysis. **(C)** Joint analysis results for PTSD case-control male analysis.

**Extended Data Figure 3.**
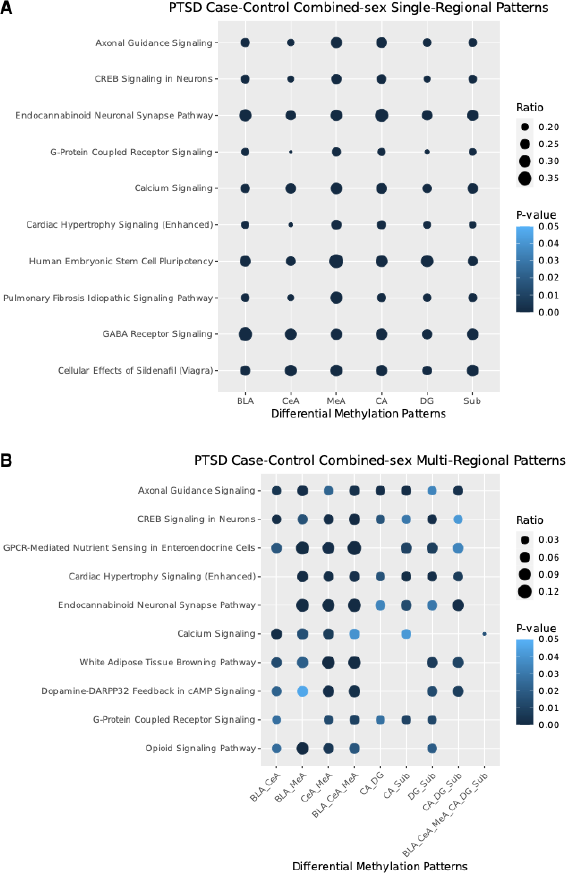
Brain region joint analysis of PTSD DNAm reveals gene set enrichment. **(A)** Canonical pathways for single-regional patterns in PTSD case-control combined-sex joint analysis. CpG sites in six single-regional differential methylation patterns were annotated to genes and Ingenuity Pathway Analysis was performed on the mapped genes (genes not mapped to the IPA database were excluded in our pathway analysis). The top 10 canonical pathways across six patterns are reported. The complete list of canonical pathways for each single-regional patterns can be found in **Supplementary Table 4. (B)** Canonical pathways for multi-regional patterns in PTSD case-control combined-sex joint analysis.

**Extended Data Figure 4.**
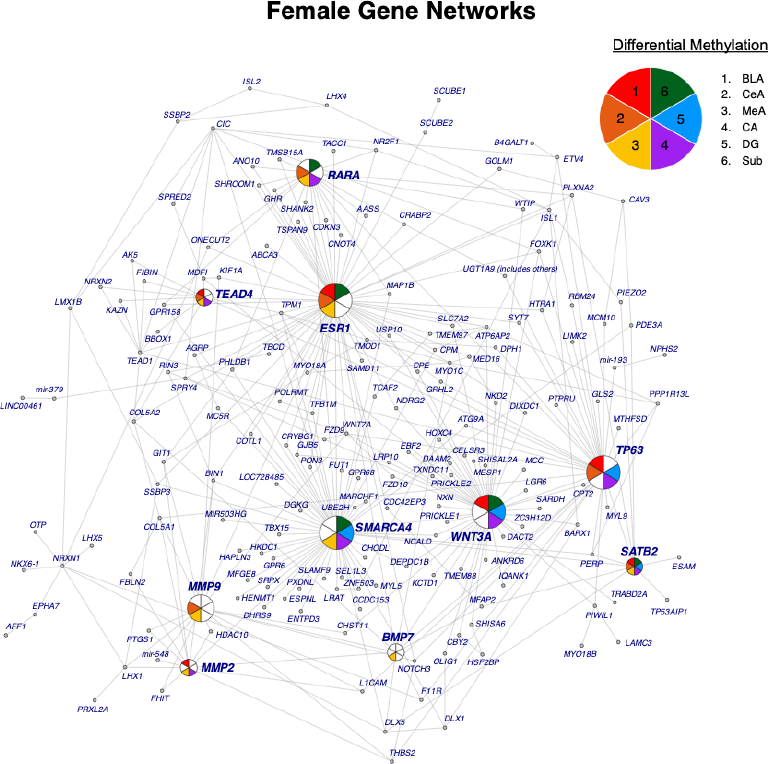
Female-specific Network analysis. Gene network analysis was performed using IPA for JMCs in each female-specific differential methylation pattern. The most significant gene networks (network score > 30) from the six single-regional analyses were obtained and merged by integrating the hub genes and their nearest neighbors in IPA. Pie-circles were used to indicate the differential methylation regions for genes that have more than two neighboring genes. The amygdala regions are colored as follows: (BLA: red, CeA: orange, and MeA: yellow) and the three hippocampus regions (CA: purple, DG: blue, and Sub: green).

**Extended Data Figure 5.**
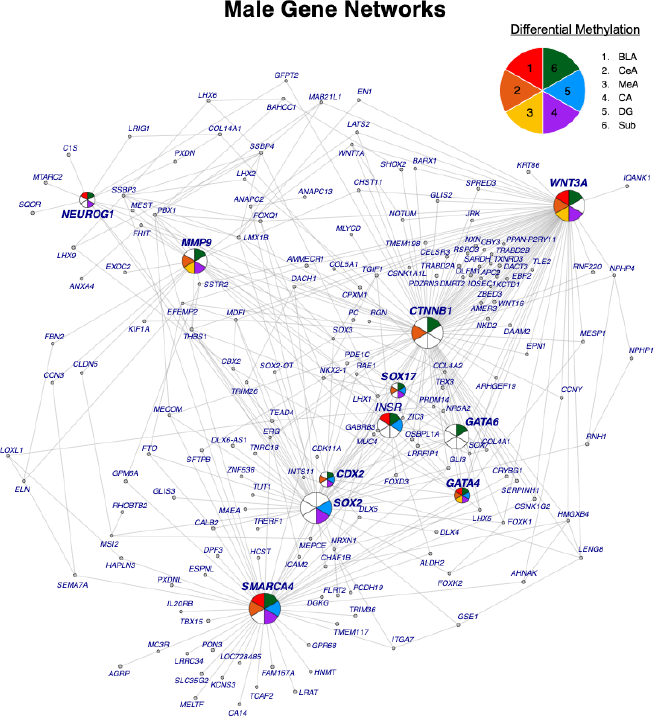
Male-specific Network analysis. Gene network analysis was performed using IPA for JMCs in each male-specific differential methylation pattern. The most significant gene networks (network score > 30) from the six single-regional analyses were obtained and merged by integrating the hub genes and their nearest neighbors in IPA. Pie-circles were used to indicate the differential methylation regions for genes that have more than two neighboring genes. The amygdala regions are colored BLA: red, CeA: orange, and MeA: yellow and the three hippocampus regions are colored CA: purple, DG: blue, and Sub: green.

**Extended Data Figure 6.**
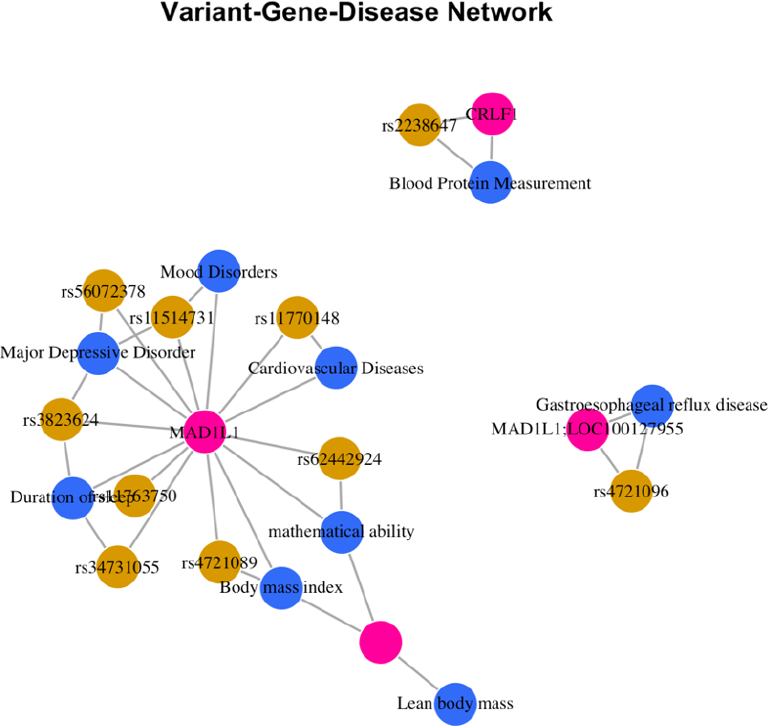
PTSD DMC GWAS Enrichment. DMCs were mapped to SNPs within the MVP GWAS to test association of DMCs with disease variants in the DisGeNet database. A variant-gene-disease network centered on MAD1L1 was identified and its risk variants and its connections to major depression and cardiovascular disease were identified.

**Extended Data Figure 7.**
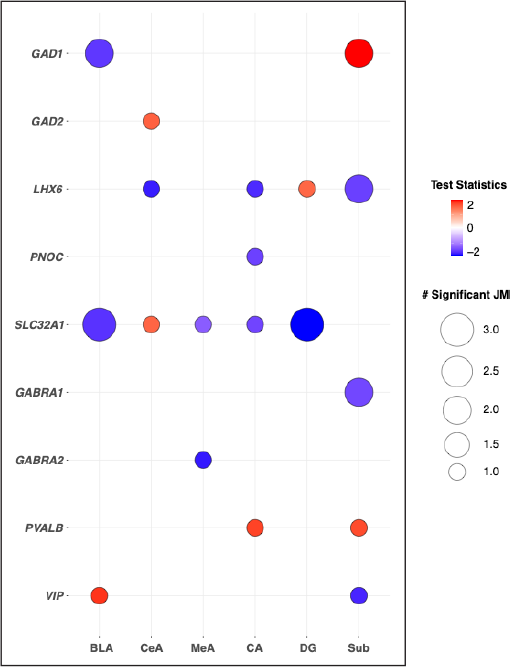
DNAm changes at interneuron marker genes. Significant enrichment of DNAm changes were also found in key drivers identified in the cortical interneuron module. Significant differential methylation for several GABAergic markers including SLC32A1, GAD1, GAD2, LHX6, PNOC, GABRA1, GABRA1, PVALB, and VIP, which were all found to be significant transcriptomic molecular drivers in this network.

**Extended Data Figure 8.**
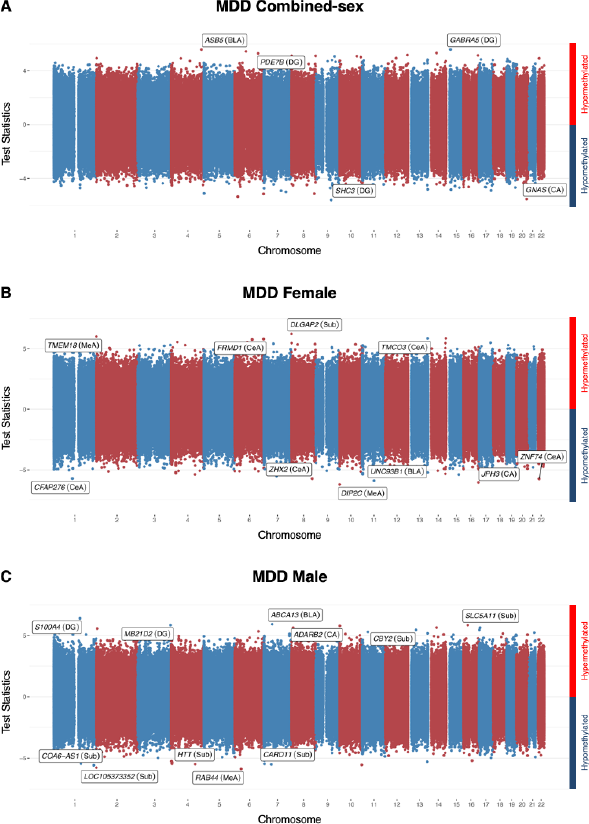
Univariate analysis reveals regional and sex-specific differences in CpG methylation between MDD cases and controls. **(A)** Manhattan plot for MDD case-control differential methylation using all samples. Differential methylation analysis was performed using a Bayesian hierarchical model, covarying for age, sex, ancestry, post-mortem interval, data source, and smoking status. The association test statistics for each variant tested is reported on the y-axis for hypermethylation (above) and hypomethylation (below). The top FDR significant markers across six regions are annotated. Labels indicate gene and regions for significant DNAm. **(B)** Manhattan plot for MDD case-control differential methylation using female samples. **(C)** Manhattan plot for MDD case-control differential methylation using male samples.

**Extended Data Figure 9.**
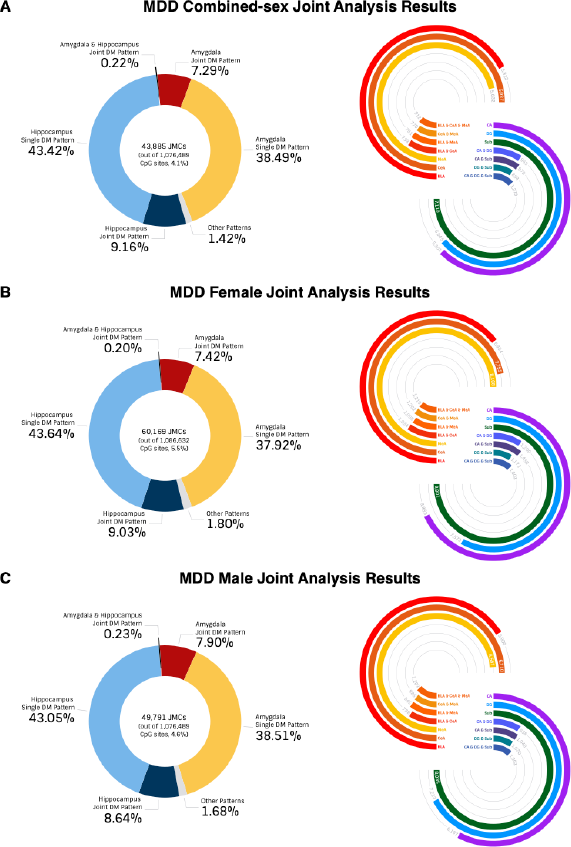
Joint analysis results for MDD case-control combined-sex analysis. **(A)** Joint analysis was performed using a Markov Random Field model incorporating summary statistics from genome-wide differential methylation analysis and topology information among brain regions. The CpG sites are classified into 64 methylation patterns. The complete list of differential methylation patterns for each CpG site can be found in **Supplementary Table 7. (B)** Joint analysis results for MDD case-control female analysis. **(C)** Joint analysis results for MDD case-control male analysis.

**Extended Data Figure 10.**
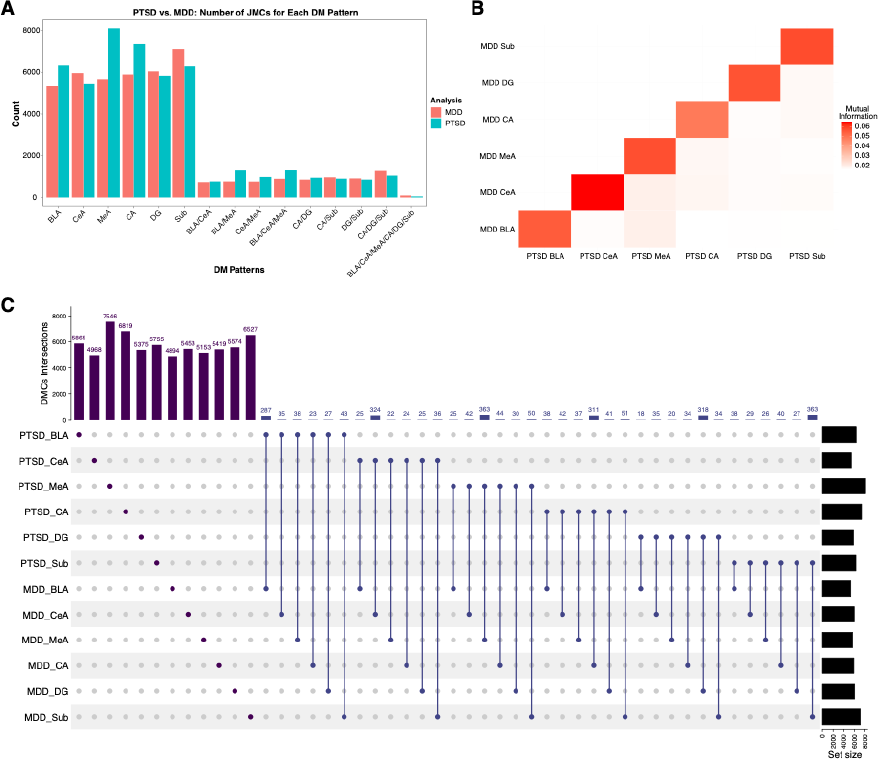
Comparison of MDD and PTSD DNAm patterns. **(A)** Bar plot comparing number of JMCs discovered between PTSD case-control analysis and MDD case-control analysis for each differential methylation pattern. **(B)** Mutual information among different analysis results across brain regions and diagnosis groups revel that between disorders, JMCs were most similar between the same regions than in any other regional or multi-regional comparison. **(C)** UpSet plot shows overlap genes for JMCs across brain regions and diagnosis groups.

## Supplementary Data Table Legends

**Supplementary Table 1.** The region-specific top 200 significant DMCs comparing each brain region versus other five regions combined.

**Supplementary Table 2.** The region-specific FDR significant DMCs from PTSD case-control combined-sex and sex-specific genome-wide differential methylation analyses.

**Supplementary Table 3.** The significant JMCs for each differential methylation pattern from PTSD case-control, combined-sex and sex-specific cross-region joint analyses.

**Supplementary Table 4.** The gene set enrichment results for combined-sex, female-specific and male-specific analyses.

**Supplementary Table 5.** The gene network results for combined-sex, female-specific and male-specific analyses.

**Supplementary Table 6.** Significant DMCs annotated to MAD1L1, ELFN1, and WNT5A genes in the ketamine response blood methylation data.

**Supplementary Table 7.** The region-specific FDR significant DMCs from MDD case-control combined-sex and sex-specific genome-wide differential methylation analyses.

**Supplementary Table 8.** The significant JMCs for each differential methylation pattern from MDD case-control combined-sex and sex-specific cross-region joint analyses.

